# Limited Role for Antibiotics in COVID-19: Scarce Evidence of Bacterial Coinfection

**DOI:** 10.1101/2020.06.16.20133181

**Authors:** Wenjing Wei, Jessica K. Ortwine, Norman S. Mang, Christopher Joseph, Brenton C. Hall, Bonnie C. Prokesch

## Abstract

**Background:** There is currently a paucity of data describing bacterial coinfections, related antibiotic prescribing patterns, and the potential role of antimicrobial stewardship in the care of patients infected with SARS-CoV-2.

**Methods:** This prospective, observational study was conducted from March 10, 2020 to April 21, 2020 in admitted patients with confirmed COVID-19. Patients were included if ≥ 18 years old and admitted to the hospital for further treatment. Data was collected via chart review from the enterprise electronic health record database. Data collected include factors driving antibiotic choice, indication, and duration of therapy as well as microbiological data.

**Findings:** Antibiotics were initiated on admission in 87/147 (59%) patients. Of these, 85/87 (98%) prescriptions were empiric. The most common indication for empiric antibiotics was concern for community-acquired pneumonia (76/85, 89%) with the most prescribed antibiotics being ceftriaxone and azithromycin. The median duration of antibiotic therapy was two days (interquartile range 1-5). No patients had a community-acquired bacterial respiratory coinfection, but 10/147 (7%) of patients were found to have concurrent bacterial infections from a non-respiratory source, and one patient was diagnosed with active pulmonary tuberculosis at the time of admission for COVID-19.

**Interpretation:** Bacterial coinfection in patients with COVID-19 was infrequent. Antibiotics are likely unnecessary in patients with mild symptoms. There is little role for broad-spectrum antibiotics to empirically treat multidrug resistant organisms in patients with COVID-19, regardless of disease severity. Antimicrobial stewardship remains important in patients infected with SARS-CoV-2.

**Funding:** The authors received no funding for this work.

## Introduction

In December 2019, a novel coronavirus, severe acute respiratory syndrome coronavirus 2 (SARS-CoV-2) was first detected in Wuhan, China and found to cause acute respiratory symptoms and pneumonia. The disease caused by SARS-CoV-2 was named coronavirus disease 2019 (COVID-19). SARS-CoV-2 has led to a global pandemic affecting over 200 countries.^1^ In the United States, cases continue to increase with over one million confirmed infections and 73,000 associated deaths as of May 2020.^2^

Patients with COVID-19 present with a variety of signs and symptoms but the majority exhibit fever, dry cough, and fatigue. Many patients also experience shortness of breath, myalgias, and anorexia amongst other less common symptoms. Disease severity can range from asymptomatic or relatively mild to severe with an estimated 20% of patients requiring admission to an intensive care unit (ICU).^3^ Chest imaging of patients with COVID-19 typically reveals bilateral multi-focal opacities on plain radiographs and bilateral, peripheral interstitial ground glass opacities on computerized tomography (CT).^2,3^ These findings are nonspecific and overlap with other infectious etiologies, creating uncertainty in differentiating COVID-19 from other common viral or bacterial respiratory infections. Thus, if bacterial pneumonia or sepsis is strongly suspected, initiation of empiric antibiotics to cover for community-acquired pneumonia (CAP) has been recommended by national guidelines.^3,4^

Bacterial infections occur both concomitantly and subsequent to a variety of viral respiratory illnesses. In the pre-antibiotic era of the 1918 influenza pandemic, bacterial infections complicated nearly all influenza-related deaths. More recently during the 2009 influenza A (H1N1) pandemic, bacterial infections were identified in up to 34% of ICU managed patients.^5^ In a typical, non-pandemic influenza season, nearly 20% of patients are diagnosed with community-acquired bacterial infections, most commonly caused by *Staphylococcus aureus* and *Streptococcus pneumoniae*.^5,6^ However, there is currently a paucity of data describing bacterial infections and related antibiotic prescribing in patients with COVID-19.

The continued development of antimicrobial resistance globally may be exacerbated in the setting of an infectious pandemic. Thus, in light of the rising number of COVID-19 cases worldwide, we believe that it is of utmost importance to continue promoting the judicious use of anti-infective agents and highlight the role of antimicrobial stewardship. The goal of this study is to assess how often patients with SARS-CoV-2 infection have clear evidence of concurrent bacterial infections and to better characterize the factors driving antibiotic prescribing, selection, and duration of therapy in this cohort of patients. This information is critical to defining the role of antimicrobial stewardship in assisting with antibiotic de-escalation and discontinuation in the management of patients with COVID-19.

## Methods

### Study design and participants

This prospective, observational study was conducted at Parkland Health & Hospital System and included patients admitted between March 10, 2020 and April 21, 2020. Parkland is an 862-bed safety net hospital as well as the primary teaching site for the University of Texas Southwestern (UTSW) Medical School providing care to underserved residents of Dallas County in Dallas, Texas and averages over one million patient visits annually. The study was approved by the UTSW Medical Center institutional review board and informed consent was waived. Patients were included if they tested positive for SARS-CoV-2 by polymerase chain reaction (PCR), were 18 years of age or older, and were admitted to the hospital for management of COVID-19. Patients were excluded if the index admission for COVID-19 was at an outside facility.

### Data collection

Patient charts were retrospectively reviewed and data was collected from the enterprise electronic health record database by the primary investigator and study personnel. Baseline characteristics collected include demographic information, significant comorbidities, smoking history, history of intravenous (IV) antibiotic exposure in the 90 days prior to admission, and COVID-19 disease severity. In addition, data regarding fever, white blood cell (WBC) count, oxygen requirement, pulmonary imaging findings, pathogen-directed infectious work up, requirement of mechanical ventilation, vasopressors, continuous renal replacement therapy, length of stay, infection with *Clostridioides difficile* during admission, and in-hospital mortality related to COVID-19 were collected. Antibiotics initiated within 48 hours of admission were recorded along with rationale, therapeutic indication, and duration of use. Antibiotics that were initiated greater than 48 hours after time of admission were considered treatment for a possible secondary bacterial infection, rather than coinfection upon admission, and thus were excluded. Clinical data and outcomes were monitored through June 1^st^,, 2020.

### Laboratory procedures

From March 10, 2020 to March 27, 2020, patients were confirmed to have SARS-CoV-2 via PCR testing on nasopharyngeal and oropharyngeal samples through outside testing facilities. On March 27, 2020, Parkland instituted in-house PCR testing on nasopharyngeal samples via the Xpert Xpress SARS-CoV-2 test manufactured by Cepheid®. Influenza/respiratory syncytial virus (RSV) PCR as well as a composite respiratory pathogen PCR panel were performed using nasopharyngeal samples. Other infectious work-up included *Legionella* urinary antigen testing and methicillin-resistant *Staphylococcus aureus* (MRSA) surveillance collected from the nares.

### Outcomes

Patients who received antibiotics on admission were compared to those who did not in order to characterize the factors driving antimicrobial prescribing in patients presenting with COVID-19. In addition, patients were assessed for evidence of community-acquired bacterial respiratory coinfection (CABRC) as well as concurrent bacterial infections from a non-respiratory source on admission.

### Definitions

The severity of COVID-19 was defined using an institution-specific management algorithm (See Supplementary Material). Fever was defined as greater than 100· 4 °F (38 °C). Leukopenia and leukocytosis were defined as a WBC count less than 4,000 cells/μL or greater than 11,000 cells/μL, respectively. CABRC was defined as presence of a positive bacterial culture consistent with CAP within 48 hours of admission and clinical signs and symptoms consistent with CAP as documented by the treatment team. Concurrent bacterial infection was defined as a positive non-respiratory bacterial culture within 48 hours of admission plus documentation consistent with active infection.

### Statistical analysis

Continuous measurements were presented as means and standard deviations (SD) or medians and interquartile ranges (IQR) and evaluated using Student’s *t-*test, or Mann-Whitney *U* test, respectively. Categorical variables were presented as counts (%) and evaluated using a *χ*2 test or Fisher’s exact test.

### Role of funding source

This study had no funder. The corresponding author had full access to all the data in the study and had final responsibility for the decision to submit for publication.

## Results

A total of 151 patients met inclusion criteria with 147 ultimately included in the study. Four patients were excluded due to having their index admission for COVID-19 at an outside facility. The average age of patients was 52 years and 60/147 (41%) were female (Table 1). One or more comorbidities were found in 114/147 patients (78%), with 16 patients (11%) having underlying chronic lung disease and 21 patients (14%) reporting being active cigarette smokers. Patients were admitted to the hospital after experiencing a median of 5 days (interquartile range, 3 to 7) of symptoms with the majority presenting with moderate severity COVID-19 disease (109/147, 74%). Seventeen patients (12%) met the criteria for severe COVID-19 with 10 (7%) requiring either mechanical ventilation or vasopressor support on admission.

**Table 1.** Clinical Characteristics of Patients and Relation to Antibiotic Prescribing

| Characteristic | Total<br>(N=147) | Antibiotic<br>(N=87) | No Antibiotic<br>(N=60) | P-value |
| --- | --- | --- | --- | --- |
| Age, years | 52 (18) | 53 (18) | 48 (15) | 0·015 |
| Female sex | 60 (41) | 34 (39) | 26 (43) | 0·606 |
| Comorbidities |  |  |  |  |
| None | 33 (22) | 23 (26) | 10 (16) | 0·231 |
| Chronic lung disease | 16 (11) | 9 (10) | 7 (12) | 0·800 |
| Hypertension | 63 (43) | 37 (43) | 26 (43) | 0·923 |
| Diabetes | 48 (33) | 25 (29) | 23 (38) | 0·223 |
| Hyperlipidemia | 35 (24) | 19 (22) | 16 (27) | 0·499 |
| Chronic kidney disease/ESRD | 14 (10) | 7 (8) | 7 (12) | 0·462 |
| Active cancer | 9 (6) | 8 (9) | 1 (2) | 0·083 |
| Congestive heart failure | 9 (6) | 3 (3) | 6 (10) | 0·160 |
| Coronary heart disease | 8 (5) | 5 (6) | 3 (5) | 1·000 |
| HIV | 5 (3) | 3 (3) | 2 (3) | 1·000 |
| Cirrhosis | 2 (1) | 1 (1) | 1 (2) | 1·000 |
| Active smoker | 21 (14) | 12 (14) | 9 (15) | 0·837 |
| IV antibiotics in the last 90 days | 4 (3) | 4 (5) | 0 (0) | 0·145 |
| COVID-19 disease severity |  |  |  |  |
| Mild | 21 (14) | 5 (6) | 16 (27) | 0·00037 |
| Moderate | 109 (74) | 65 (75) | 43 (73) | 0·681 |
| Severe | 17 (12) | 17 (20) | 0(0) | 0·00027 |
| Presentation |  |  |  |  |
| Pneumonia on imaging | 124 (84) | 79 (91) | 45 (75) | 0·010 |
| Fever on admission | 105 (71) | 66 (76) | 39 (65) | 0·152 |
| Fever > 3 days | 71 (48) | 45 (52) | 26 (43) | 0·317 |
| Leukopenia | 20 (14) | 10 (11) | 10 (17) | 0·369 |
| Leukocytosis | 12 (8) | 12 (14) | 0 (0) | 0·004 |
| Supplemental oxygenation | 80 (54) | 55 (63) | 25 (42) | 0·010 |
| Infectious labs ordered |  |  |  |  |
| None | 22 (15) | 4 (5) | 18 (30) | <0·0001 |
| Influenza/RSV | 39 (27) | 33 (38) | 6 (10) | 0·00016 |
| Respiratory viral panel<sup>a</sup> | 36 (24) | 30 (34) | 6 (10) | 0·00069 |
| *Legionella* urine antigen | 51 (35) | 41 (47) | 10 (17) | 0·00014 |
| Respiratory culture | 47 (32) | 36 (41) | 11 (18) | 0·0032 |
| Blood culture | 112 (76) | 77 (89) | 35 (58) | < 0·0001 |
| Urine culture | 45 (31) | 31 (36) | 14 (23) | 0·112 |
| MRSA nares screen | 19 (13) | 18 (21) | 1 (2) | 0·00073 |
| Community-acquired bacterial respiratory infection | 0 (0) | 0 (0) | 0 (0) | · |
| Concurrent bacterial infection from non-respiratory source | 10 (7) | 10 (11) | 0 (0) | 0·0056 |
| Tuberculosis | 1 (1) | 1 (1) | 0 (0) | 1·00 |
| Mechanical ventilation at anytime – n/total n(%)<sup>†</sup> | 29/141 (21) | 23/81 (28) | 6/60 (10) | 0·0138 |
| CRRT at anytime– n/total n(%)<sup>†</sup> | 5/141 (4) | 4/81 (5) | 1/60 (2) | 0·394 |
| Vasopressor support at anytime– n/total n(%)<sup>†</sup> | 22/141 (16) | 18/81 (22) | 4/60 (7) | 0·0175 |
| LOS > 7 | 64 (44) | 45 (52) | 19 (32) | 0·025 |
| CDI | 0 (0) | 0 (0) | 0 (0) | · |
| In-hospital Mortality – n/total n(%)<sup>†</sup> | 13/141 (9) | 9/81 (11) | 4/60 (6) | 0·398 |

Data presented as n (%) or mean (SD) unless otherwise specified
Abbreviations: CDI, *Clostridioides difficile* infection; ESRD, end stage renal disease; HIV, human immunodeficiency virus; LOS, length of stay
<sup>a</sup>Respiratory viral panel tests for Adenovirus, Coronavirus 229E, Coronavirus HKU1, Coronavirus NL63, Coronavirus OC43, Influenza A, Influenza A/H3, Influenza A/2009-H1, Influenza B, Human Metapneumovirus, Human Rhinovirus/Enterovirus, Parainfluenza 1-4, RSV, *Bordetella pertussis*, *Chlamydomphila pneumoniae*, and *Mycoplasma pneumoniae*
<sup>†</sup>As of June 1<sup>st</sup>, 2020, 6 patients remain admitted in the hospital, leaving 141 patients able to be assessed for mortality

Antibiotics were initiated within 48 hours of admission in 87/147 patients (59%). Of these, 85 patients (98%) received antibiotics as empiric therapy, and 2 (2%) had antibiotics continued from an outpatient course. The most common indication for empiric antibiotics (Table 2) was CAP (76/85, 89%). The median duration of antibiotic therapy for any indication was 2 days (interquartile range, 1 to 5). Following the introduction of in-house PCR testing for SARS-CoV-2, a shorter duration of antibiotic therapy was noted (Figure 1). The majority of patients (74/85, 87%) were exposed to two or more antibiotics during the empiric course of therapy, most commonly ceftriaxone and azithromycin (Table 3). Broad-spectrum antibiotic therapy (vancomycin, piperacillin/tazobactam, and/or cefepime) was prescribed in only 24/147 (16%) patients. Of these 24 patients, only four patients had a recent history of IV antibiotic exposure in the 90 days prior to admission and none had a history of MRSA or *Pseudomonas aeruginosa*.

**Table 2.** Indications for Empirically Prescribed Antibiotics

| **Empirically Selected Indication** | **Number of Patients (%)<sup>a</sup>** |
| --- | --- |
| Community acquired pneumonia | 76 (90) |
| Urinary Tract Infection | 5 (6) |
| Skin and Soft Tissue Infection | 2 (2) |
| Neutropenic Fever | 1 (1) |
| Meningitis | 1 (1) |
| Central Line-associated Bloodstream Infection | 1 (1) |
| Chorioamnionitis | 1 (1) |
| Colitis | 1 (1) |
| Spontaneous Bacterial Peritonitis | 1 (1) |

<sup>a</sup>Patients could have more than one indication

**Table 3.** Empiric Antibiotics Prescribed

| Antibiotic | Number of Prescriptions |
| --- | --- |
| Ceftriaxone | 68 |
| Azithromycin | 57 |
| Vancomycin | 22 |
| Doxycycline | 17 |
| Piperacillin/tazobactam | 16 |
| Cefepime | 4 |
| Moxifloxacin | 3 |
| Ampicillin | 3 |
| Gentamicin | 2 |
| Clindamycin | 2 |
| Amoxicillin | 1 |
| Metronidazole | 1 |
| Trimethoprim/sulfamethoxazole | 1 |
| Ciprofloxacin | 1 |
| Amoxicillin/clavulanate | 1 |

**Figure 1.**
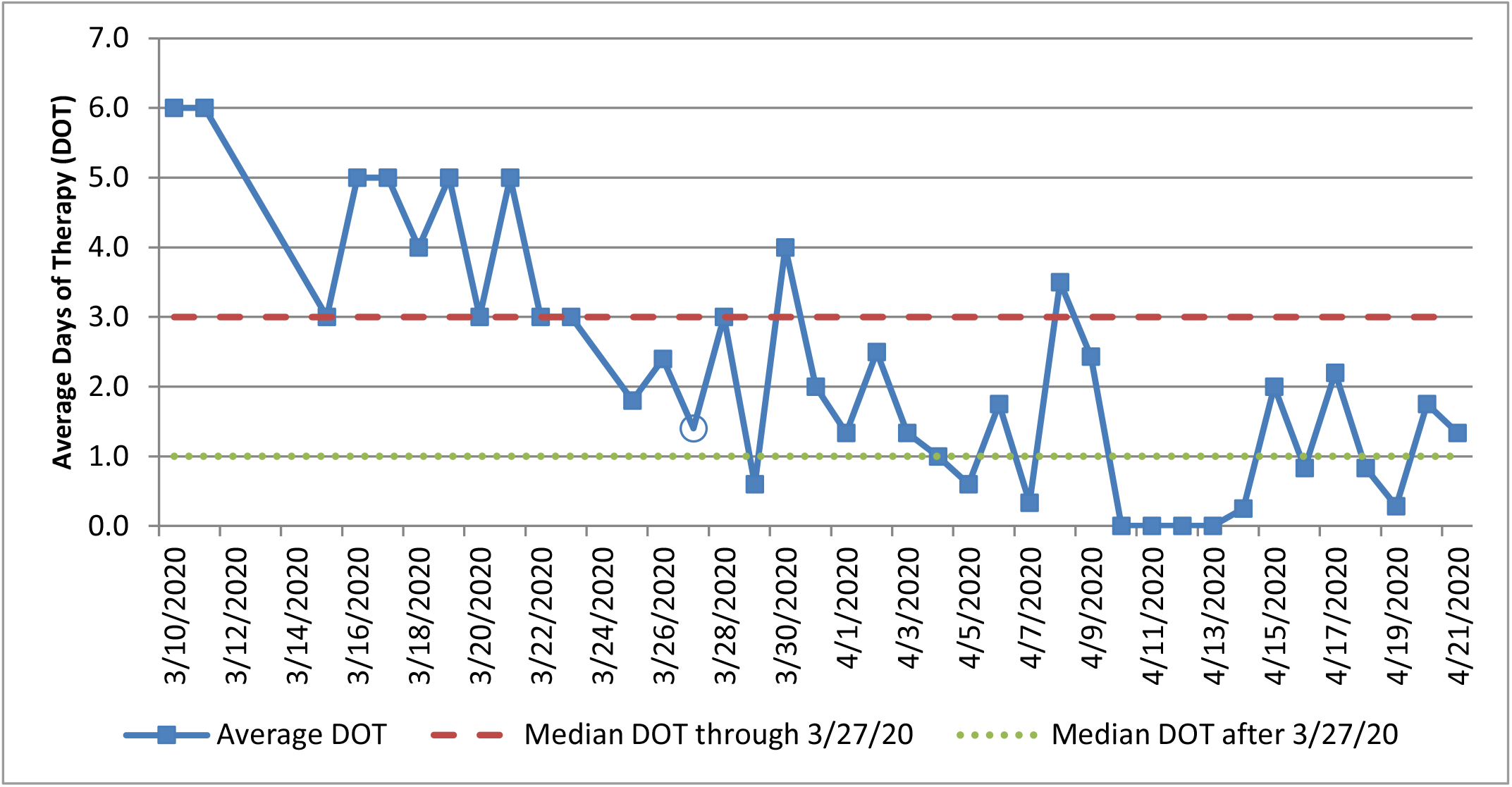
Average Duration of Therapy Based on Patient Admission Date; 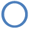, date of implementation of in-house SARS-CoV-2 PCR testing.

Antibiotic prescribing was significantly more common in patients with severe disease, evidence of pneumonia on imaging, leukocytosis, or supplemental oxygen requirements on admission. Patients who presented with mild disease were significantly less likely to receive antibiotics. There was no difference in frequency of antibiotic prescribing in patients with moderate disease or in those who were febrile on admission. Patients initiated on antibiotics upon admission also underwent a significantly more robust infectious workup than those who were not started on empiric antibiotic therapy (Table 1).

While respiratory cultures were ordered on 47/147 (32%) patients, none returned positive for significant bacterial growth. All Legionella urine antigen tests were negative. Most patients had blood cultures drawn on admission (112/147 [76%]), including all 24 patients who were started on broad-spectrum antibiotics. In addition, 45/147 (31%) had urine cultures sent and 19/147 (13%) were screened for MRSA nares colonization. No proven CABRCs were identified in our patient cohort. However, clinical suspicion remained high enough that 19/76 (25%) patients who received empiric antibiotics for CAP on admission completed at least five days of therapy. Overall, 10/147 (7%) of patients were found to have concurrent bacterial infections unrelated to a respiratory source and one patient was diagnosed with active pulmonary tuberculosis (Table 4). Although nine patients were found to have positive blood cultures on admission, eight cultures were deemed contaminants (see Supplementary Material). One patient was considered to have a veritable bacteremia which was secondary to a gastrointestinal source. Similarly, ten patients had positive urine cultures on admission, but only five were considered pathogenic per treatment team documentation.

**Table 4.** Concurrent Bacterial Infections From a Non-Respiratory Source

| **Concurrent Bacterial Infection** | **Patients (N=10)** |
| --- | --- |
| Urinary tract infection | 5 (3) |
| Skin and soft tissue infection | 2 (1) |
| Bacteremia | 1 (1) |
| Otitis media | 1 (1) |
| Chorioaminionitis | 1 (1) |

Data presented as n (%)

## Discussion

Early epidemiological studies of patients with COVID-19 reported empiric antibiotic use in 71-100% of patients, with Chen and colleagues reporting a median duration of therapy of five days (interquartile range, 3 to 7).^7–11^ Comparatively, we found lower rates of empiric antibiotic utilization with shorter durations of therapy overall. Longer durations of therapy were noted earlier in the outbreak and may be correlated with lengthy turnaround times (more than one week) to receive final SARS-CoV-2 testing results from a commercial reference laboratory. Once in-house testing was established, turnaround times decreased dramatically with results typically available to clinicians within two hours. This likely contributed to increased levels of physician comfort in withholding empiric antibiotics in more stable patients given the prompt return of diagnostic testing, as well as facilitating more rapid antibiotic de-escalation in those patients testing positive for COVID-19. The availability of rapid on-site testing for SARS-CoV-2 plays an important role in the decision-making process for discontinuation of antibiotic therapy.

Antibiotic choice was not reported in most prior studies published on this topic, but empiric agents primarily targeted common CAP pathogens. Wang and colleagues reviewed antibiotic use among 102 patients with COVID-19 and observed 87 (85%) patients received quinolones, 34 (33%) cephalosporins, and 25 (25%) carbapenems, while Cao and colleagues reported receipt of moxifloxacin in 39/67 (58%) patients and antifungal therapy in 8/67 (12%).^11,12^ Unlike previously published literature which showed a high use of quinolones and carbapenems, we observed more narrow-spectrum antibiotic utilization. This is consistent with the 2019 American Thoracic Society/Infectious Diseases Society of America (ATS/IDSA) practice guidelines for CAP, which recommend combination therapy with an IV beta-lactam (e.g. ceftriaxone) plus azithromycin for patients admitted with CAP in the absence of risk factors for infections caused by multi-drug resistant organisms.^13^

We observed limited use of broad-spectrum agents in general, though that may be because this study focused only on antibiotics prescribed within 48 hours of hospital admission. The ATS/IDSA CAP guidelines recommend empirically treating MRSA or *Pseudomonas aeruginosa* only if specific risk factors are present. These risk factors include recent hospitalization with receipt of IV antibiotics, prior history of either pathogen in the last 12 months, or high local prevalence rates for either pathogen.^13^ Only four patients in this study had risk factors for multi-drug resistant (MDR) organisms, and therefore the majority of patients were appropriately prescribed narrow-spectrum antibiotics. Most of the patients initiated on broad-spectrum regimens were de-escalated quickly if MRSA surveillance screen and/or blood cultures were negative. Based on these observations, we recommend that careful assessment of MDR risk factors be performed before initiating broad-spectrum antibiotics and cultures should be obtained to help guide de-escalation. MRSA nasal screening has a negative predictive value of > 95% for MRSA pneumonia. The utilization of MRSA surveillance screening to assist with early de-escalation should be encouraged in order to decrease unnecessary exposure to vancomycin, lab draws and monitoring, and reduce risk of nephrotoxicity.^14,15^

There is currently limited information available regarding rates of bacterial coinfections with COVID-19. However, bacterial coinfection rates of 0-47% and 2-65% were reported in systematic reviews of pandemic influenza H1N1 and of influenza and other respiratory viruses, respectively.^16,17^ While *S. pneumoniae* was the most commonly identified organism, MRSA and nosocomial Gram-negative organisms were also reported. Differences in illness severity, timing of infection, and whether coinfection was documented on admission or resulted as a complication of prolonged hospital stay, mechanical ventilation, or secondary to the virus may have contributed to the variability in reported rates. In a prospective analysis of CAP by Abelenda-Alonso and colleagues only 57/1123 (5·1%) patients had influenza and a bacterial coinfection on admission, which is similar to the minimal evidence of coinfection in our study.^18^ Because COVID-19 has emerged recently, there is limited literature regarding bacterial coinfections in the setting of primary SARS-CoV-2 infection, but a review of 18 studies describing bacterial coinfections in patients with any coronavirus infection was performed by Rawson and colleagues.^19^ The authors described low rates of bacterial coinfection among the nine studies published for COVID-19 (62/806 [8%]). However, most studies were not specifically evaluating coinfections and thus did not report the organisms identified. The low rates of bacterial coinfections among patients with respiratory viral illnesses, including COVID-19, are similar to the findings in our cohort of 147 patients. Interestingly, the average time to development of a bacterial superinfection in patients with influenza has been reported to be 7-14 days after the onset of the viral infection.^20^ Therefore, the fact that none of the patients in our cohort were found to have definitive evidence of bacterial coinfection on admission is not unusual, as the patients presented a median of five days from symptom onset.

The median duration of antibiotic therapy in our cohort was short, indicating that suspicion for bacterial coinfection was low with only 19/147 (13%) patients receiving five days or more of empiric antibiotic therapy for CAP. Due to concern for increased infection transmission, most respiratory samples were collected from throat swabs rather than sputum or lower respiratory tract samples and half of the samples obtained from sputum were rejected due to being unsatisfactory quality specimens. Although identification of organisms may have been limited by this inability to obtain quality respiratory cultures, sputum cultures overall have poor yield for pathogen isolation. Our institution does not perform *S. pneumoniae* urine antigen testing; however, both *S. pneumoniae* and *Legionella* urine antigen tests have modest sensitivity for clinical disease and the most recent ATS/IDSA CAP guidelines do not recommend routinely testing these urine antigens in adults with non-severe CAP.^14^ Procalcitonin has been suggested as a potentially useful biomarker to differentiate bacterial and viral infections and assist with antibiotic decision-making.^21–23^ However, due to the lack of data regarding its reliability in completely ruling out bacterial pneumonia with accuracy, the role of procalcitonin in COVID-19 is currently unknown.^13,24^ Although blood cultures are not routinely recommended in non-severe CAP, they were collected from a majority of patients in this cohort. Blood culture results were ultimately not helpful in identifying clinically significant pathogens as nearly all organismal growth was considered to be from skin contamination. Therefore, blood cultures are likely not necessary in patients presenting with mild to moderate COVID-19 who do not meet the criteria for severe CAP.

In summary, we identified zero cases of CABRC in patients with COVID-19. While it is reasonable to initiate empiric antibiotics for possible bacterial infection in clinically severe patients awaiting diagnostic confirmation of COVID-19, broad-spectrum agents are likely unnecessary in the absence of risk factors for MDR organisms. Based on this study, it appears antibiotics are of limited utility in the setting of proven COVID pneumonia. If antibiotics are initiated, they should be de-escalated early in patients positive for SARS-CoV-2 with no other evidence of bacterial infection within 48 hours. Antimicrobial stewardship has an important role in limiting unnecessary antibiotic exposure and optimizing resources during this COVID pandemic.

## Contributors

WW, JKO, NSM, and BCP conceived and designed the study. WW and CJ performed the data collection. WW and BCH performed data analysis. WW drafted the manuscript and all authors participated in critical revision of the manuscript for important intellectual content. All authors approved the final manuscript and were responsible for the decision to submit for publication.

## Data Availability

All data in the manuscript is contained within the manuscript or supplements submitted.

## Declaration of Interests

The authors report no relevant conflicts of interest.

## Notes

### Competing Interest Statement

The authors have declared no competing interest.

### Funding Statement

The authors received no external funding for this work.

### Author Declarations

This study was approved by the IRB at the University of Texas Southwestern Medical Center and site approval was given by Parkland Health and Hospital System.

